# Opioids, Needle Exchange, and Barriers to Care in the Rural Context: Interviews with Law Enforcement and Emergency Medical Personnel

**DOI:** 10.1101/2021.04.28.21256269

**Authors:** Ralph Lawton, John Leland, William Leland

**Author notes:** Correspondence Ralph Lawton, 419 Chapel Drive, 213 Social Sciences Building, Box 90097, Durham, NC, 27708.

## Abstract

**Background:** The large increase in opioid use, subsequent addiction and related death has come to be known as a national epidemic in the United States, particularly affecting rural areas. The crisis has been met with an increase in attempts to address the large overdose rates. In many of these strategies, first responders such as police and emergency services personnel are placed on the front lines. The input and opinions of these individuals, and the communication with them from agency leadership during program implementation can dramatically affect the success of drug related policies. However, there is a paucity of research focused on the attitudes of primary responders, particularly in rural settings. This study conducts a geographically-focused investigation into the attitudes of rural EMS and law enforcement personnel in regards to opioid treatment in a location that was currently implementing a major substance use policy.

**Methods:** Qualitative semi-structured interviews were conducted with 24 members of law enforcement and 22 emergency medical providers within a single rural county in North Carolina. Interviews spanned three sections: demographics, barriers to opioid treatment and county-specific programs.

**Results:** The largest barriers to care cited were lack of local treatment resources, the stigma against drug use and a perception that people who use opioids did not want to change. A multi-agency approach to the crisis was supported by EMS personnel and law enforcement. However, first responders’ awareness of an active multiagency county-wide initiative was very limited. This was very surprising, as first responders were expected to implement the program, and this phenomena resulted in very low program efficacy to that point. This result underscored the importance of communication between and within agencies, particularly in environments that are resource-constrained.

**Conclusions:** This study is unique in comparing attitudes of rural EMS and law enforcement regarding substance use treatment. This paper provides insight into the viewpoints of rural first responders, with clear implications for rural drug policy. This paper further underscores opportunities for maximizing effective opioid policy in rural settings, particularly emphasizing clear communication between agencies.

## 1. Introduction

In the past two decades, the United States has seen a massive increase in both opioid prescriptions and in deaths from opioid and opiate overdose, including from heroin. The large increases in opioid prescriptions and the related deaths from overdoses have led to what has been described as an “epidemic” of drug overdose deaths, with a nearly 200% increase in overdose deaths involving opioids since 2000[1, 2].

North Carolina in particular saw a 73% increase in opioid deaths from 2005-2015 and saw a 40% increase in opioid deaths from 2015-2016 alone[3]. The more rural, eastern parts of North Carolina have been particularly intensely affected. Some counties in Eastern North Carolina reported a tripling in the number of opioid deaths between 2011-2015, as well as a sharper increase in general overdoses than recorded statewide[4, 5].

The surge in overdoses has been met with an increase in attempts to combat the rising tide, many of which place first responders – such as police and emergency services personnel – on the front lines. Particularly, initiatives to equip police and emergency services with naloxone have begun to pull these organizations into the coordinated public health efforts to address addiction and overdose[6, 7]. During overdose emergencies, law enforcement and EMS are often both dispatched, and, over the course of routine community work, both groups frequently work with individuals who use opioids. The input and opinions of these individuals can dramatically affect the successful implementation of drug-related policies and therefore has significant policy implications. A long body of evidence has underscored the importance of first responder opinions within law enforcement, and recent evidence highlights the importance of first responders and their opinions as keys in proposed drug-related initiatives[8-12].

There is a paucity of published data focused on the attitudes of primary responders in a rural setting. Given the dramatic differences that specific contexts can make on the quality, availability and delivery of care, our study focuses on the opinions and attitudes of first responders within a single county in rural Eastern North Carolina regarding the challenges faced dealing with substance use. The particular county where interviews were focused is currently in the midst of the rollout of several coordinated programs with their local health department, sheriff’s office, EMS and fire department in order to tackle an escalating overdose crisis. This research draws on the critical perspectives of people on the front lines, those whose daily work bridges care between treatment organizations and individuals with substance use disorders.

## 2. Methods

### 2.1. Participants

Data were collected between 2017-2018 in a totally rural county in Eastern North Carolina[13]. The county also has some of the highest poverty levels in the state, with over 25% of people living below the poverty line. Researchers interviewed 24 members of county law enforcement and 22 EMS personnel who encountered people who used opioids in their day-to-day work. Participants were recruited using a snowball sampling technique, beginning with health department contacts in each group and followed by recommendations from EMS and law enforcement members. In-person interviews were conducted on five different days at each site in an effort to mitigate selection of specific individuals that may have been present in a single time period. Written informed consent was obtained prior to each interview, which lasted approximately 20 minutes. Participants were not paid to participate.

Police and EMS member interviews were digitally recorded, but their personal information was anonymized for analysis and reporting.

### 2.2. Data Collection

This was a data-driven, inductive qualitative study. A qualitative semi-structured interview protocol was used for each group.

Law enforcement and emergency personnel interviews had three sections: demographics, barriers to opioid treatment and county programs. Data were collected on each individual’s gender, age, marital status, children, race, education, income and role in their respective organization.

Participants were also asked about barriers to opioid treatment in the area. They were asked an open-ended question to list what major barriers to care they observed in the county.

Interviewees were also asked of their knowledge on a specific multi-agency program being rolled out by the county to address the opioid epidemic. They were asked questions regarding their knowledge on the existence of the program, whether they believed it would be successful and if they thought it could be more effective if hosted in an alternate setting.

### 2.3 Analysis

Using constant comparison techniques, we constructed primary descriptive codes and subcodes. Coding took place concurrently with interviews, so that even as interviews were being conducted, researchers were coding the data and discussing amongst each other between interviews. A code structure was determined as codes emerged. For example, if an EMS responder were to cite “lack of public transportation to treatment facilities,” and another were to discuss “long distances of travel to treatment,” these were combined with other transportation-related issues into the code “lack of transportation.” Once a stable code list was established (Table 1), codes were applied to responses independently by two reviewers per transcript, followed by an intercoder agreement exercise to reach consensus on any disagreement. If there were any disagreements, a third reviewer was consulted. Interviews of EMS and police department personnel were stopped when data saturation was reached[14]. By the end of interviews in both pools, no new codes had emerged in the last series of interviews, and the researchers determined that no new nuances of existing codes were being revealed. [Table 1]

**Table 1:** Themes coded for each “Domain” investigated in the interviews.

| <u>Domain</u> | <u>Themes</u> |
| --- | --- |
| **<u>Barriers to Care</u>** | Lack of Resources |
|  | Lack of Personal Support |
|  | Stigma in small-population setting |
|  | Apathy |
|  | Poor Communication |
| **<u>Treatment Programs</u>** | Accessibility |
|  | Poor Communication |

### 2.4 Ethics Approval

This study protocol was approved by the Duke University Institutional Review Board (ref. Number 2018-0006).

## 3. Results

<u>Overall, EMS and law enforcement personnel agreed on most topics concerning the opioid crisis and how to address it.</u>

### 3.1. Demographics

Summary statistics on demographic data in each group are presented in Table 2. Law enforcement was older & more male than EMS. Over 20% more members of law enforcement were married. A nearly identical proportion were White, and similar proportions had postsecondary education. Significantly more members of law enforcement had four-year degrees.

**Table 2:** Demographics of EMS personnel and law enforcement.

|  | EMS | Law Enforcement |
| --- | --- | --- |
| Median Age | 33 | 39 |
| % Male | 45.5% | 83.3% |
| % Married | 40.9% | 62.5% |
| % White | 72.7% | 70.8% |
| % with any postsecondary ed. | 63.6% | 74.2% |
| % with 4-year degree | 4.54% | 29.2% |
| Personal/Family Experience | 33% | 62.5% |
| Total # | 22 | 24 |

**Table 3:** Code List for Barriers to Care Barriers to Care.

|  |
| --- |
| **<u>Barriers to Care</u>** |
| Lack of Treatment Resources |
| Stigma |
| Willpower |
| Lack of Knowledge |
| Lack of Transportation |
| Dealing with mental health issues<br>they're not trained in |
| High Costs |

According to the census data, the study county is approximately 45% White, 45% Black and 10% other races. While not surprising given that White populations tend to be overrepresented in law enforcement, EMS and law enforcement groups were both approximately 70% White and are not very representative of the county population. However, our samples match the reported demographics of each group as a whole. The data on age and gender for the law enforcement personnel generally matched the national averages for these values (39.6 years old and 87% male respectively), though the EMS personnel were slightly younger and more female[15]. [Table 2]

We also collected data on whether or not the EMS personnel and law enforcement members had personal or familial experience with opioid misuse. Law enforcement had personal experience much more frequently than EMS.

### 3.2. Barriers to Care

#### Transportation difficulties compound a lack of resources

When asked about barriers to care, a majority of both EMS and law enforcement personnel cited lack of local treatment resources as a key concern. The nearest substance use treatment center is outside of the county, and most of the psychiatric care in the area is difficult to access without insurance. This lack of access was noted by EMS providers and law enforcement personnel in the course of their work. In parallel with these concerns about the lack of local resources were statements discussing that lack of transportation is a common problem, making out-of-county resources even more inaccessible. One EMS member said there was a complete lack of resources for people who use drugs:

> *“Nothing comes to my mind when I think about getting help in (this) County.”*

Another EMS worker expressed this lack of support felt:

> *“They don’t feel they have people to reach out to.”*

A law enforcement officer said there isn’t anything nearby where people who use opioids can receive help:

> *“There’s simply nothing here. It’s easier to get heroin than Methadone from the nearest clinic.”*

#### In the rural setting, stigma plays an outsized role in decisions to seek care

A large portion of EMS personnel and law enforcement also discussed the role of stigma in making treatment difficult. They felt that, particularly in a lower-population rural area, stigmas can play an outsized role in shaping behaviors, dissuading many individuals from seeking treatment. One law enforcement member specifically discussed the effects of the rural setting on seeking treatment:

> *“Everyone knows what everyone is up to around here, I wouldn’t go to an exchange in the middle of town.”*

#### First responders hold a prevailing belief in lack of willpower amongst patients

Concerns that the “lack of willpower” among individuals with substance misuse disorders were keeping them from seeking treatment were very common. Particularly, there seemed to be a common perception that people who use opioids simply do not want to stop, or are at least apathetic.

One EMS member said opioid addiction often seems impossible to overcome:

> *“It seems like a hole they can never get out of.”*

Another EMS worker said people who use drugs often make major sacrifices to obtain opioids:

> *“It doesn’t matter what you have to do or who you have to hurt. You have to get that high.”*

Other EMS workers thought that there was an apathetic element standing in the way of effective treatment:

> *“I don’t feel like they care. They’re looking for the next hit.”*

#### Perceived associations between treatment and law enforcement can complicate access

Particularly amongst law enforcement, people often expressed concerns that people with substance misuse disorders would perceive legal ramifications to seeking care. As will be discussed in depth later, this is of particular relevance to the opioid-treatment placement programs that the county has undertaken. One law enforcement member said people who use opioids are often skeptical of the sheriff’s department:

> *“They all don’t trust there won’t be legal ramifications. They think if they don’t complete the program to our standards, we’ll put a target on their back.”*

### 3.4 Attitudes regarding opioid treatment

EMS and law enforcement personnel were split on the most effective ways to treat the opioid epidemic in the county. About two years prior, the county had implemented a multiagency program (including EMS, county health & human services, and law enforcement) housed at the sheriff’s office in which individuals with opioid use disorders would be able to turn in their opioids without concern of conviction and receive assistance and placement into treatment. Having seen successful programs elsewhere, the leadership in these agencies were highly hopeful that a similar system would work well in their county.

#### A majority of first responders supported a multiagency approach, though concerns with apathy and stigma re-arose

When asked about a hypothetical program that would accomplish the same goals as the county program that already existed, both EMS and members of law enforcement were generally supportive, although many thought the program being housed in the police department would deter people who use opioids from seeking help. One EMS worker doubted that a person who used drugs would seek help from the police:

> *“You really think a person on drugs will go to the sheriff’s office? Nope.”*

Another expressed the same sentiment:

> *“I just can’t imagine a drug dealer going to the police department. Sorry.”*

As with care-seeking more generally, worries about apathy amongst people with opioid misuse disorders resurfaced, as did concerns about stigma. One EMS member doubted that people who use opioids would seek help at all:

> *“They would use it if they wanted help.”*

Despite the concerns about locating the program in the police department, over two-thirds of each group expressed support for this general sort of program and thought it would be effective. One member of law enforcement emphasized this view:

> *“Given how little is here, and how hard it is to find care, any additional ways that we can help people stop using and find some treatment are a good thing, even if we have to do it in the police department.”*

#### First responders emphasized the role of providing information to individuals with opioid misuse disorders

A common thread through many of the interviews was the importance of disseminating knowledge, and making sure information regarding treatment and prevention reached individuals with opioid misuse disorders. One law enforcement member emphasized this role of education and information in finding care:

> *“The most important thing is education, if they know where to find treatment, then the ones that really want it will go to it.”*

#### Despite broad support for the idea, most first responders were unaware of their agency’s own programs

Despite the emphasis on education and information, when asked about the existing county program by name, the majority of EMS and law enforcement personnel were not aware of it. As a result, they had not been promoting it to the target populations. This was highly surprising, as the first responders interviewed were supposed to be responsible for implementing the program.

## 4. Discussion

In this study, we conducted a semi-structured series of interviews of law enforcement and EMS personnel, focused within a single rural county in Eastern North Carolina. This enabled us to do an inductive assessment on the barriers to care in this context and the challenges programs in these settings may face, and to discuss opinions of opioid treatment programs - in particular a flagship interagency program run by the county. First responders broadly shared similar views on topics concerning the opioid crisis. One notable difference was the high rate of personal experience with drug misuse among law enforcement. Given previous literature on law enforcement burnout, this difference underscores the personal nature and prevalence of the issue, and the implications of proper management for first-responder wellbeing[16]. However, the reason why the rates were higher among law enforcement is unclear.

Although substance use disorders and overdoses have become larger public health concerns, it is often still difficult to receive proper care and overcome barriers to appropriate healthcare in rural areas. Major focus has been placed on the rural-urban divide in both the burden of the issue and the different obstacles that exist in these places. Specifically, urban areas tend to provide more auxiliary services that are necessary for successful outcomes, and they tend to have a more diverse and convenient array of treatment options[17-19]. This is in line with what we found in a rural setting. The top-cited barrier in both groups to care was the lack of resources to deliver treatment. This lack of resources was discussed as a major contributor to the general sense of frustration from EMS personnel. Given the established disparities in inpatient services, auxiliary care and psychiatrists in rural areas, this is not surprising. Further underscoring this disparity in opioid treatment was that, while the local health care facility was able to stabilize an overdose, comprehensive management could require 40-minute ambulance rides with high acuity patients to a fully capable hospital.

Other features inherent to rural areas pose challenges to receiving treatment. Small populations and large family networks can lead to issues with confidentiality and stigma, which may discourage people with mental health or opioid use disorders from seeking care or resources[20]. The data we have collected is consistent with this hypothesis, as both interviewed groups suggested stigma was a major concern with regards to receiving care. Beyond the concerns with stigma, there were broader concerns about apathy among people who use opioids and lack of familial or community support. Taken together, these major concerns with the lack of resources, stigma and transportation all suggest that as limited resources get deployed, it will be of utmost importance to ensure that they are as physically accessible and confidential as possible. The concerns with communication and dissemination of information are critical, and the results we find highlight the need for effective and coordinated communication within and between organizations for programs to be effective. It may be the case as well that effective communication of community efforts may reduce feelings of apathy among people who use opioids. However, it is clear from the repeated discussion of the lack of addiction and mental health treatment resources that additional investment in these facilities is required to deal with this problem in the long term.

Another major concern that arose was communication with regards to the area’s flagship program. While multiple agencies came together to roll out a coordinated program to try and help facilitate treatment, this knowledge ultimately did not make it down to the rank-and-file of each group, including the department housing the initiative. This was very surprising, as the success of the program ultimately depended on the implementation by first responders. First responders’ lack of knowledge of state policies has been documented before and can be somewhat expected given the detached nature in which state policy often gets implemented[21]. However, we believe this to be the first instance documenting first responder lack of knowledge of their own flagship programs. While county-wide programs like that offered in the county studied have been implemented before in other locations[22], those programs often only consist of needle exchange and typically are well-received and publicized. This underscores the need not just for inter-agency cooperation, but effective communication of new programs throughout the entirety of each agency as well.

## 5. Conclusion

While the opioid epidemic affects people everywhere, the effects may be particularly harsh in rural areas which are underserved and understudied, and where the treatment options for caregivers and people who use opioids are limited. This study is unique in comparing attitudes and opinions of rural EMS and law enforcement regarding substance use treatment. The opinions of these first responders highlights the many barriers to care and underscores the importance of a holistic approach to treatment which addresses issues such as transportation, confidentiality and minimizing stigma. Our case study of a specific county dramatically underscores the importance of effective inter and intra-agency communication. We conclude that significant additional investment in mental health and addiction treatment resources is necessary, and that accessibility and confidentiality should be at the forefront of designing these programs. We also underscore the importance of effective law enforcement collaboration and buy-in, suggesting that focusing on needlestick concerns may make law enforcement more willing to collaborate. Lastly, we strongly emphasize the importance of effective inter and intra-agency communication in implementation of treatment programs.

## Data Availability

Data is available upon request

## Acknowledgements

We acknowledge Karen LaChapelle, Wilson Muse, and John Britt for their assistance in data collection. We acknowledge Duncan Thomas, PhD, for assistance designing the project. We acknowledge Paul Seale, MD, and Paul Mihas, PhD, for their consulting on the project.

## References

1. Dart RC, Surratt HL, Cicero TJ, Parrino MW, Severtson SG, Bucher-Bartelson B, Green JL. Trends in opioid analgesic abuse and mortality in the United States. The New England Journal of Medicine. 2015; 372(3): 241–248. https://doi.org/10.1056/nejmsa1406143 PMid: 25587948

2. Rudd RA, Aleshire N, Zibbell JE, Gladden RM. Increases in drug and opioid overdose deaths—United States, 2000–2014. American Journal of Transplantation. 2016; 16(4): 1323–1327. https://doi.org/10.15585/mmwr.mm6450a3 PMid: 26720857

3. Kansagra SM, Cohen MK. The Opioid Epidemic in NC Progress, Challenges, and Opportunities. North Carolina medical journal. 2018; 79(3): 157–162. https://doi.org/10.18043/ncm.79.3.157 PMid: 29735617

4. Harper A. Opioid Crisis Strikes Area. Rocky Mount Telegram. Accessed (September 12, 2018).http://www.rockymounttelegram.com/Tarboro/2017/09/12/Opioid-crisis-strike-Edgecombe-County.html

5. North Carolina Association of County Commissioners. Opioid Abuse Statistics by County. 2017. https://www.ncacc.org/648/Opioid-Abuse-Statistics-by-County

6. Kanouse AB, Compton P. The epidemic of prescription opioid abuse, the subsequent rising prevalence of heroin use, and the federal response. Journal of pain & palliative care pharmacotherapy. 2015; 29(2): 102–114. https://doi.org/10.3109/15360288.2015.1037521 PMid: 26095479

7. Nicolaidis C. Police officer, deal-maker, or health care provider? Moving to a patient-centered framework for chronic opioid management. Pain Medicine. 2011; 12(6): 890–897. https://doi.org/10.1111/j.1526-4637.2011.01117.x PMid: 21539703

8. Beletsky L, Macalino GE, Burris S. Attitudes of police officers towards syringe access, occupational needle-sticks, and drug use: a qualitative study of one city police department in the United States. International Journal of drug policy. 2005; 16(4), 267–274. https://doi.org/10.1016/j.drugpo.2005.01.009

9. Beyer L, Crofts N, Reid G. Drug offending and criminal justice responses: practitioners’ perspectives. International Journal of drug policy. 2002; 13(3): 203–211. https://doi.org/10.1016/S0955-3959(02)00063-4

10. Marinos V, Innocente N. Factors influencing police attitudes towards extrajudicial measures under the youth criminal justice act. Canadian Journal of Criminology and Criminal Justice. 2008 Jul; 50(4): 469–489. https://doi.org/10.3138/cjccj.50.4.469

11. Petrocelli M, Oberweis T, Smith MR, Petrocelli J. Assessing police attitudes toward drugs and drug enforcement. American Journal of Criminal Justice. 2013; 39(1): 22–40.

12. Tobin KE, Gaasch WR, Clarke C, MacKenzie E, Latkin CA. Attitudes of emergency medical service providers toward naloxone distribution programs. Journal of Urban Health. 2005; 82(2): 296–302. https://doi.org/10.1093/jurban/jti052 PMid: 15917504

13. Health Resources and Services Administration. Federal Office of Rural Health Policy - Defining Rural Population. 2018; hrsa.gov/rural-health/about-us/definition/index.html

14. Urquhart C. Grounded Theory for Qualitative Research: A Practical Guide. Thousand Oaks: Sage. 2013;

15. Cross, C. L., & Ashley, L.. Police trauma and addiction: Coping with the dangers of the job. FBI Law Enforcement Bulletin. 2004; 73, 24.

16. Data USA. Emergency Care Attendants and Law Enforcement from American Community Survey Public Use Microdata Sample. 2016; datausa.io/profile/cip/510810/

17. Keyes KM, Cerdá M, Brady JE, Havens JR, Galea S. Understanding the rural–urban differences in nonmedical prescription opioid use and abuse in the United States. American journal of public health. 2014; 104(2): e52–e59. https://doi.org/10.2105/AJPH.2013.301709 PMid: 24328642

18. Pullen E, Oser C. Barriers to substance abuse treatment in rural and urban communities: Counselor perspectives. Substance use & misuse. 2014; 49(7): 891–901. https://doi.org/10.3109/10826084.2014.891615 PMid: 24611820

19. Schoeneberger ML, Leukefeld, CG, Hiller ML, Godlaski T. Substance abuse among rural and very rural drug users at treatment entry. The American Journal of Drug and Alcohol Abuse. 2009; 32(1): 87–110. https://doi.org/10.1080/00952990500328687 PMid: 16450645

20. Sowell RL, Lowenstein A, Moneyham L, Demi A, Mizuno Y, Seals, BF. Resources, stigma, and patterns of disclosure in rural women with HIV infection. Public Health Nursing. 2007; 14(5): 302–312. https://doi.org/10.1111/j.1525-1446.1997.tb00379.x PMid: 9342922

21. Banta-Green CJ, Beletsky L, Schoeppe JA, Coffin PO, Kuszler PC. Police officers’ and paramedics’ experiences with overdose and their knowledge and opinions of Washington State’s drug overdose–naloxone–Good Samaritan law. Journal of Urban Health. 2013; 90(6): 1102–1111. https://doi.org/10.1007/s11524-013-9814-y PMid: 23900788

22. Davis SM, Davidov D, Kristjansson AL, Zullig K, Baus A, Fisher M. Qualitative case study of needle exchange programs in the Central Appalachian region of the United States. PloS One. 2018; 13(10): e0205466. https://doi.org/10.1371/journal.pone.0205466 PMid: 30312333

